# Genomic insights into the population structure and recent expansion of *Coccidioides* in the United States

**DOI:** 10.64898/2026.07.17.26358348

**Authors:** Emanuel M. Fonseca, Kevin Perry, Bridget M. Barker, Megan Hirschi, Kimberly E. Hanson, Katharine S. Walter

**Author notes:** **Corresponding Author**: Emanuel M. Fonseca.

## Abstract

**Background:** Coccidioidomycosis is an emerging fungal disease across the arid Americas and a frequent cause of community-acquired pneumonia. Understanding where *Coccidioides* populations originate, how they move across space, and whether they are expanding is important for interpreting changing patterns of Valley fever and anticipating future infection risk.

**Methods:** We prospectively collected and whole-genome sequenced 186 *Coccidioides*-positive clinical isolates submitted to a national diagnostic laboratory, and included 126 previously sequenced genomes. We applied genomic clustering, time-calibrated phylogenetic reconstruction, ancestral area reconstruction, mating-type assignment, and demographic inference to identify major populations, infer dispersal patterns, assess evidence for recombination and clonality, and reconstruct historical population dynamics.

**Findings:** We analyzed 312 genomes (139 *C. immitis*; 173 *C. posadasii*) and identified three major genetic populations within each species. *C. immitis* included two California-centered populations and one Pacific Northwest population, whereas *C. posadasii* included two Arizona-centered populations and one Texas-centered population. The most recent common ancestor was estimated at approximately 127,000 years for *C. immitis* and 234,000 years for *C. posadasii*. Most populations were not fully monophyletic, consistent with retained ancestral variation and/or ongoing gene flow. Inferred dispersal was largely asymmetric, with most movement originating from California in *C. immitis* and from Arizona and Texas in *C. posadasii*. Most populations contained both mating types, but one *C. immitis* population and a Brazilian subgroup of *C. posadasii* were clonal. All populations showed recent demographic expansion.

**Interpretation:** The evolutionary history of *Coccidioides* is characterized by strong geographic structure, ongoing gene flow, and recent demographic expansion. These processes are likely to influence future patterns of Valley fever endemicity and supports the use of genomic surveillance to detect shifts in disease risk as environmental conditions change.

**Funding:** Burroughs Wellcome Fund; Wilkes Center for Climate Science & Policy

## Introduction

Coccidioidomycosis (Valley fever) is an environmentally acquired fungal disease found across arid regions of the Americas and rapidly increasing in incidence in the United States (U.S.)^1,2^. Infections are caused by inhalation of airborne spores from *Coccidioides immitis* and *C. posadasii*, and are a common cause of community acquired pneumonia, sometimes leading to disseminated infection and death. Reported cases in the U.S. rose nearly tenfold over the past two decades, from 2,271 cases in 1998 to more than 21,037 cases in 2023, although true incidence is estimated to be at least 10 times higher due to substantial underdiagnosis and underreporting^3^. While Arizona and California continue to account for the highest disease burden, infections are more frequently detected across a widening geographic range, including low-incidence states and regions historically considered non-endemic, such as the Pacific Northwest and Intermountain West^4,5^.

Reported case increases likely reflect multiple factors, including improved awareness, testing, and reporting^6–8^, while climate change, prolonged drought, human population growth, and land-use changes may alter environmental suitability, dust generation, and exposure risk^9–12^. Climate-based niche models predict that suitable habitat for both *C. immitis* and *C. posadasii* may expand substantially over the coming decades, potentially doubling the geographic extent of endemic regions by the end of the century and disease incidence increasing by more than 50% under high carbon-emissions scenarios^9^. These trends raise concern that coccidioidomycosis will continue to expand geographically and impose an increasing public health burden.

Reported coccidioidomycosis cases provide only a partial and indirect measure of *Coccidioides* infection risk. Case surveillance remains incomplete because coccidioidomycosis is not a reportable in all states, including the known endemic state of Texas, and many infections remain undiagnosed throughout the US^13^. Case data indicate where disease is detected, not where the fungus originates, or disperses^14,15^. Thus, key questions remain about *Coccidioides,* including the timing of emergence, the frequency and direction of movement among regions, the role of particular areas as geographic sources (“hotspots”), and the influence of historical climatic change on population demography.

Previous genomic studies have identified population structure within both *C. immitis* and *C. posadasii*^16–19^, revealed environmental reservoirs outside of traditional endemic zones^16–19^, and shown that some infections in historically non-endemic regions can be either locally acquired or travel associated^4,16,20^. These studies reveal genomic evidence of sexual recombination in *Coccidioides* despite its uncharacterized sexual cycle^19–21^. Mating-type distributions can help distinguish populations with recombination potential^17,21,22^ from those showing evidence of clonal expansion^17,22^. Clarifying how *Coccidioides* populations are spreading requires an integrated analysis of population relationships, geographic origins, dispersal history, and demographic change across broad spatial and temporal scales. Thus, the integration of newly sequenced clinical isolates from both high– and low-incidence regions with publicly available genomes from underrepresented regions of the United States and Latin America provides an opportunity to revisit *Coccidioides* evolution in a broader dataset.

Here, we prospectively collected clinical isolates from a national diagnostic laboratory and combined them with publicly available genomes to conduct a national level phylogeographic study of *Coccidioides* in the United States and Latin America. Using whole-genome sequencing and phylogeographic analyses we characterized population structure, inferred historical dispersal and demographic change, and estimated the timing of diversification in both *C. immitis* and *C. posadasii*. Together, these analyses provide a genomic framework for understanding the evolutionary history of *Coccidioides* and interpreting changing patterns of coccidioidomycosis endemicity.

## Methods

### Sample Collection

We prospectively collected *Coccidioides-*positive clinical isolates submitted to ARUP Laboratories (Table S1), a U.S. reference diagnostic laboratory, between January 2023 and November 2024. ARUP Laboratories serves as a national reference laboratory, however its client base is not evenly distributed across the United States. The geographic distribution of isolates reflects the distribution of submitting clients rather than uniform national sampling, and therefore may not directly represent the true environmental or clinical distribution of *Coccidioides*. Isolates were derived primarily from sputum, in addition to soft-tissue, skin, bone, cerebrospinal fluid, and urine. Culture and processing followed CLSI M54 guidelines^23^, and DNA was extracted from a single colony grown on Sabouraud dextrose agar. Isolates were identified at ARUP Laboratories using matrix-assisted laser desorption/ionization time-of-flight mass spectrometry (MALDI-TOF MS).

Because isolates were deidentified at the submitting hospital or laboratory, only the ZIP code of the submitting laboratory was available, rather than the patient’s residential address. This introduces two sources of geographic uncertainty: first, laboratory location does not necessarily reflect the patient’s location, and second, the place of diagnosis or specimen submission may not reflect the place of exposure or infection. Accordingly, because *Coccidioides* is an environmentally acquired pathogen, we could not assign the infection site or corresponding environmental reservoir to any individual isolate. We therefore restricted geographic analyses to isolates from endemic states (Fig. 1) and interpreted all spatial inferences with caution. Deidentified diagnostic isolates also lacked additional patient-level clinical data.

**Figure 1.**
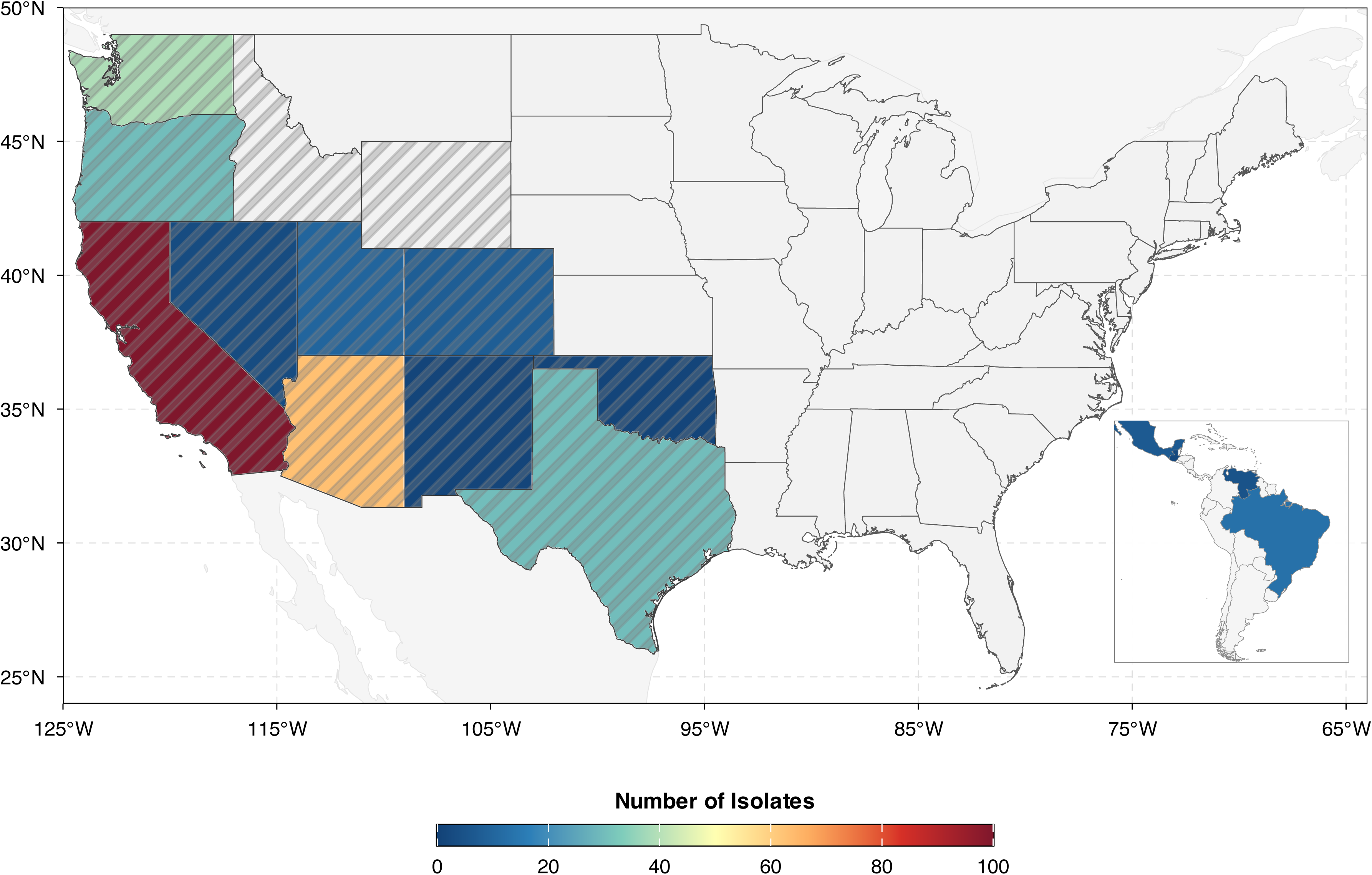
Map showing the geographic distribution of all *Coccidioides spp.* genomes included in this study across the United States and neighboring regions. Color indicates the number of isolates collected from each state or region, with darker red tones representing higher sampling density. Hatched states denote regions considered endemic for *Coccidioides*.

Following deidentification under a University of Utah Institutional Review Board (IRB)-approved protocol, we generated 186 genomes, including 89 *C. immitis* and 97 *C. posadasii*. These were combined with 126 publicly available genomes from prior studies, yielding a final dataset of 312 genomes: 139 *C. immitis* and 173 *C. posadasii*. Detailed information on all genomes included in this study is provided in Table S1, including whether each sequence was newly generated or previously available, along with accession numbers, species assignm ents, and sampling locations.

### Human Subjects

This study was approved by the University of Utah Institutional Review Board (IRB_00164551).

### Whole Genome Sequencing

We extracted genomic DNA from fungal cultures at ARUP Laboratories using the Maxwell RSC Cell DNA Purification Kit (AS1370). We enzymatically fragmented 1–25 ng of DNA and prepared libraries using the New England Biolabs NEBNext Ultra II FS DNA Library Prep Kit (cat# E7805L), targeting an average insert size of approximately 350 bp. We assessed library quality using an Agilent Technologies 4150 TapeStation with D1000 ScreenTape assays (cat# 5067-5582 and 5067-5583). We normalized and pooled libraries, then sequenced them as 2 × 150 bp paired-end on an Illumina NovaSeq X Series platform at the University of Utah High-Throughput Genomics Core.

### Variant Calling

We identified genomic variants with the cocci-call pipeline^24^. The pipeline trimmed adapters and low-quality bases (Phred < 20), screened reads with Kraken2^25^ to remove non-*Coccidioides* reads and assigned species using the log-transformed ratio of unique minimizers classified as *C. immitis* or *C. posadasii*. For alignment, cocci-call mapped reads to the appropriate species reference genome (*C. immitis* GCF_000149335.2^26,27^; *C. posadasii* GCA_018416015.2^28,29^) using BWA-MEM^30^ (v0.7.17), marked duplicate reads with GATK v4.3^31^, and masked repetitive elements using NUCmer^32^ and RepeatMasker http://www.repeatmasker.org/. We excluded samples with mean coverage < 30×. For all downstream analyses, we restricted the genomic dataset to intergenic sites identified from SnpEff annotations^33^. We used this conservative marker set to reduce the contribution of coding variation, where purifying selection, local adaptation, and linked selective processes can introduce signals that reflect functional constraint or adaptation rather than neutral population history. This filtering was intended to minimize systematic biases in population structure, branch-length estimation, molecular-clock dating, and demographic inference, although the sensitivity to departures from neutrality differs among these analyses. Addicionally, to reduce non-independence among linked regions, we applied linkage disequilibrium (LD)-pruning using a sliding window approach (10 kb window, step size of 1 variant) and retained variants at an r^2^ threshold of 0.2 using Plink^34^.

### Genetic structure

We investigated genetic structure using the sparse Non Negative Matrix Factorization (sNMF) algorithm implemented in the R package LEA^35,36^ and Discriminant Analysis of Principal Components (DAPC) in adegenet^37^. For sNMF, we ran 15 independent replicates for values of K (number of genetic clusters) ranging from 1 to 10 under a haploid model with 1,000 iterations per run. We selected K using cross-entropy and a first-plateau approach, defined as the last value before the relative improvement in mean cross-entropy dropped below 0.5%. We used DAPC as a complementary, model-free approach. We selected The optimal number of clusters using the Bayesian Information Criterion (BIC) obtained from successive K-means clustering. The suggested K was defined as the last value before the relative improvement in BIC dropped below 0.5%. We retained primary population assignments only when both approaches supported the same assignment. When the methods disagreed, we conservatively adopted the less subdivided assignment^38^.

### Phylogenetic Reconstruction

We reconstructed relationships among the sampled isolates using a maximum likelihood framework implemented in IQ-TREE 2^39^. We selected the nucleotide substitution model using ModelFinder^40^ under the Bayesian Information Criterion and applied ascertainment bias correction to account for the absence of invariant sites. We assessed node support with 1,000 ultrafast bootstrap replicates^41^. Then, we inferred time-scaled phylogenies with TreeTime^42^ by calibrating branch lengths under a molecular clock framework. We converted genetic distances into temporal divergence-time estimates using fixed substitution rates from previous calibrated Bayesian phylogenomic analyses: 1.08 × 10□□ substitutions per site per year for *C. posadasii* and 1.02 × 10□□ substitutions per site per year for *C. immitis*. These rates derive from a time-calibrated tree in which the divergence between *C. immitis* and *C. posadasii* was anchored using a fossil-based calibration derived from broader fungal phylogenies, as first established in comparative genomic analyses of *Coccidioides*^26^.

To assess sensitivity to linkage disequilibrium, we compared LD-pruned and unpruned phylogenetic reconstructions. Root-to-tip branch lengths were moderately correlated between datasets, with stronger concordance in *C. immitis* than in *C. posadasii* (Spearman’s ρ = 0.732 and 0.505, respectively; Fig. S1). LD pruning shortened and narrowed root-to-tip distances in both species, indicating that linked sites inflated branch length and divergence time estimates, especially in *C. posadasii* (Fig. S2).

### Ancestral Area Reconstruction

We inferred the geographic origins and movement of *Coccidioides* populations through time using maximum-likelihood ancestral area reconstruction with the *ace* function implemented in the R package ape^43^. We treated sampling localities as discrete characters, using the submitting diagnostic laboratory for newly sequenced isolates and the recorded U.S. state or country for publicly available genomes. The analysis included 18 localities: 15 U.S. states and three countries outside the United States. We projected ancestral state estimates onto the phylogeny to infer the most probable geographic origins of major populations and historical connections among regions^18,44,45^.

### Mating type inference

We inferred the mating type (MAT) locus of each isolate using a direct read-mapping approach^46^. We extracted reads from BAM files generated by the variant-calling pipeline and remapped them to MAT1-1 (EF512012) and MAT1-2 (XM_001246634) using BWA-MEM^47^ (v0.7.19). We sorted and indexed alignments with SAMtools^48,49^ (v1.16), then quantified the total mapped reads, mean sequencing depth, and breadth of coverage. We considered a MAT locus detected when it had ≥50 mapped reads, mean depth ≥5X, and coverage breadth ≥50%. We assigned a mating type when only one locus passed these criteria. When both loci showed evidence of mapping, we required a minimum five-fold difference in support to assign the dominant locus; Otherwise, samples were classified as ambiguous. If neither locus passed these thresholds, samples were classified as having no detectable MAT locus.

### Changes in effective population size through time

We reconstructed the demographic history of *C. posadasii* and *C. immitis* using StairwayPlot2^50^, which infers changes in effective population size from the site frequency spectrum (SFS). For each population, we generated a folded SFS to summarize minor allele frequencies without inferring the ancestral state at each site. We ran StairwayPlot2 with default optimization settings and scaled demographic reconstructions using previously estimated substitution rates^17^ of 1.08 × 10□□ substitutions per site per year for *C. posadasii* and 1.02 × 10□□ substitutions per site per year *for C. immitis*. We assumed a generation time of one year^19^. We used these parameters to convert coalescent units to estimates of effective population size (N□) and calendar time.

## Results

### Population genetic structure

Over one year of genomic surveillance, we generated 203 *Coccidioides* genomes from patients across 23 states. We excluded 17 genomes came from patients outside of recognized *Coccidioides-*endemic regions for a final surveillance dataset that included 186 genomes from endemic states, comprising 89 newly sequenced *C. immitis* and 97 *C. posadasii* isolates (Fig. 1). These isolates span the known geographic range of both species, encompassing hyperendemic regions^51^, with persistently high incidence of disease, and endemic areas with lower incidence^3,52,53^. To place these data in a broader phylogenomic context, we incorporated an additional 126 previously published *Coccidioides* genomes, yielding a combined dataset suitable for national-scale phylogeographic inference (Table S1).

Population structure analyses identified three major populations within each species. To ensure robust population assignments, we compared clustering results from sNMF and DAPC. Although the two approaches recovered broadly similar structure, they differed for some admixed individuals (Figure 2a–b). We therefore used a conservative consensus framework based on concordance between methods. In *C. immitis*, this approach identified three populations: one centered in California with additional isolates from Oregon, a second largely restricted to the Pacific Northwest, and a third distributed across California, Oregon, Washington, and Utah (Fig. 3a–c).

**Figure 2.**
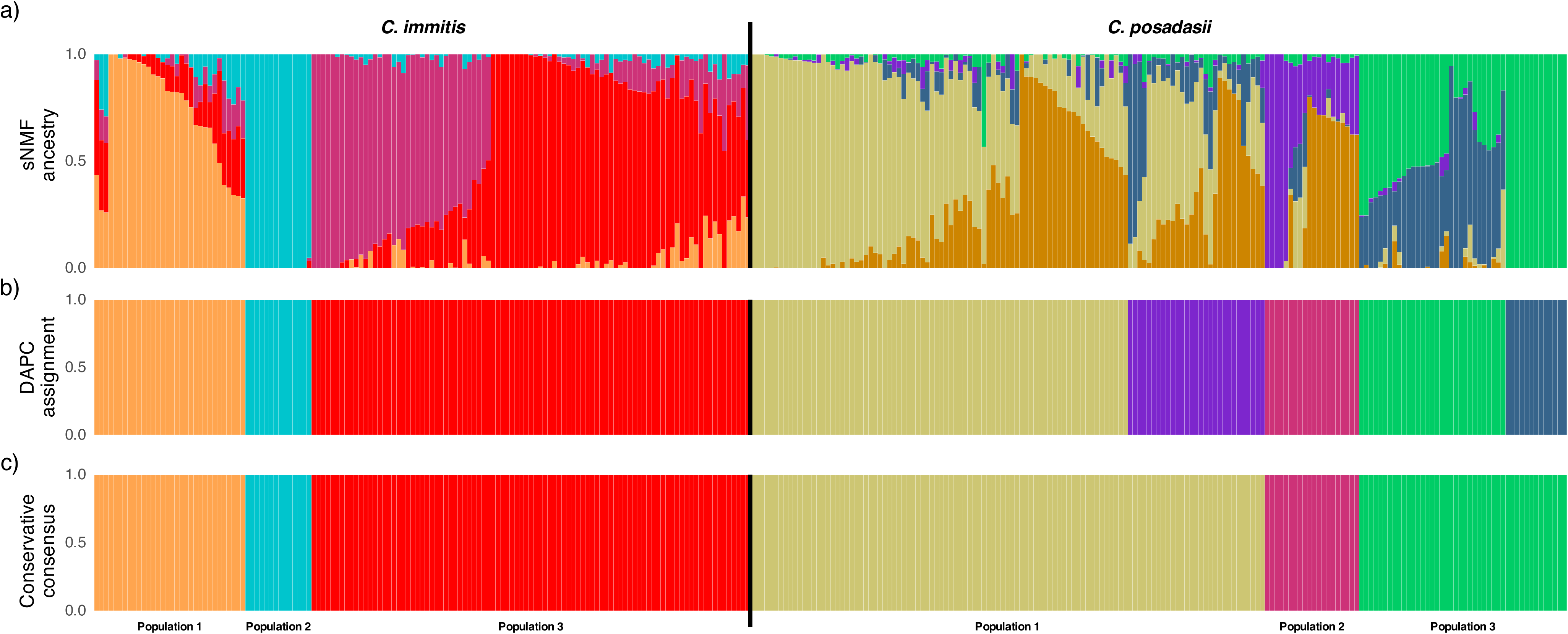
Population genetic structure inferred from genome-wide SNP data. Each vertical bar represents an individual isolate. (a) Ancestry proportions estimated using sparse non-negative matrix factorization (sNMF). Individuals with admixed ancestry are indicated by the presence of two or more colors in a single vertical bar. (b) Population assignment inferred using discriminant analysis of principal components (DAPC).

**Figure 3.**
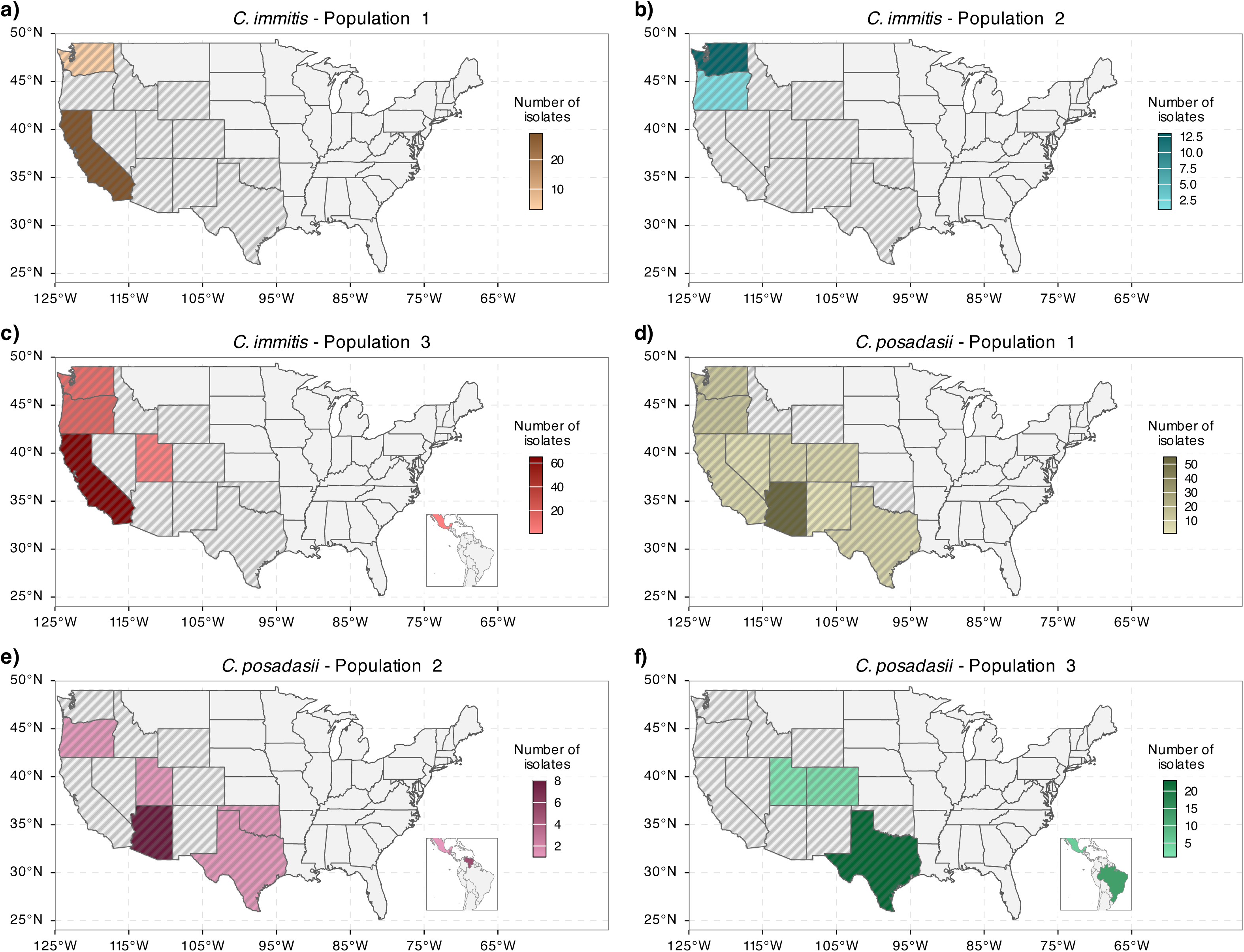
Spatial distribution of the major *Coccidioides spp.* populations across North America. Each panel shows the number and geographic distribution of isolates belonging to a given genomic population by state or region, with shading indicating the number of isolates.

In *C. posadasii*, the consensus framework also identified three populations. Population 1 was broadly distributed across the western United States, with most isolates from Arizona and additional isolates from California, Nevada, Utah, New Mexico, and Texas (Fig. 3d). Population 2 spanned Arizona, Texas, and Oregon, and extended into Central and South America through isolates from Mexico, Guatemala, and Venezuela (Fig. 3e). Population 3 was centered in Texas and included additional isolates from Utah, New Mexico, Mexico, and Brazil (Fig. 3f).

### Phylogenetic relationships and divergence timing

We reconstructed time-calibrated phylogenies for both species and estimated that the most recent common ancestor (MRCA) of sampled *C. immitis* existed approximately 127,000 years ago (95% CI: 112,000–143,100 years ago), whereas the MRCA of sampled *C. posadasii* dated to approximately 233,500 years ago (95% CI: 208,200–261,900 years ago) (Fig. 4). Major phylogenetic groupings were broadly consistent with the populations inferred from population structure analyses, although most populations were not monophyletic. Within *C. immitis*, population 2 formed a monophyletic clade with a recent MRCA of approximately 3,700 years ago (95% CI: 2,800–4,500 years ago), indicating recent population-specific diversification. In contrast, Populations 1 and 3 were not monophyletic and were consistent with the species-level MRCA of approximately 127,000 years ago (95% CI: 112,000–143,100 years ago). In *C. posadasii*, none of the three populations were monophyletic, and all traced back to ancient MRCAs of approximately 195,000–234,000 years ago. Pairwise divergence estimates were similarly ancient, suggesting deep population structure that has not resolved into distinct lineages.

**Figure 4.**
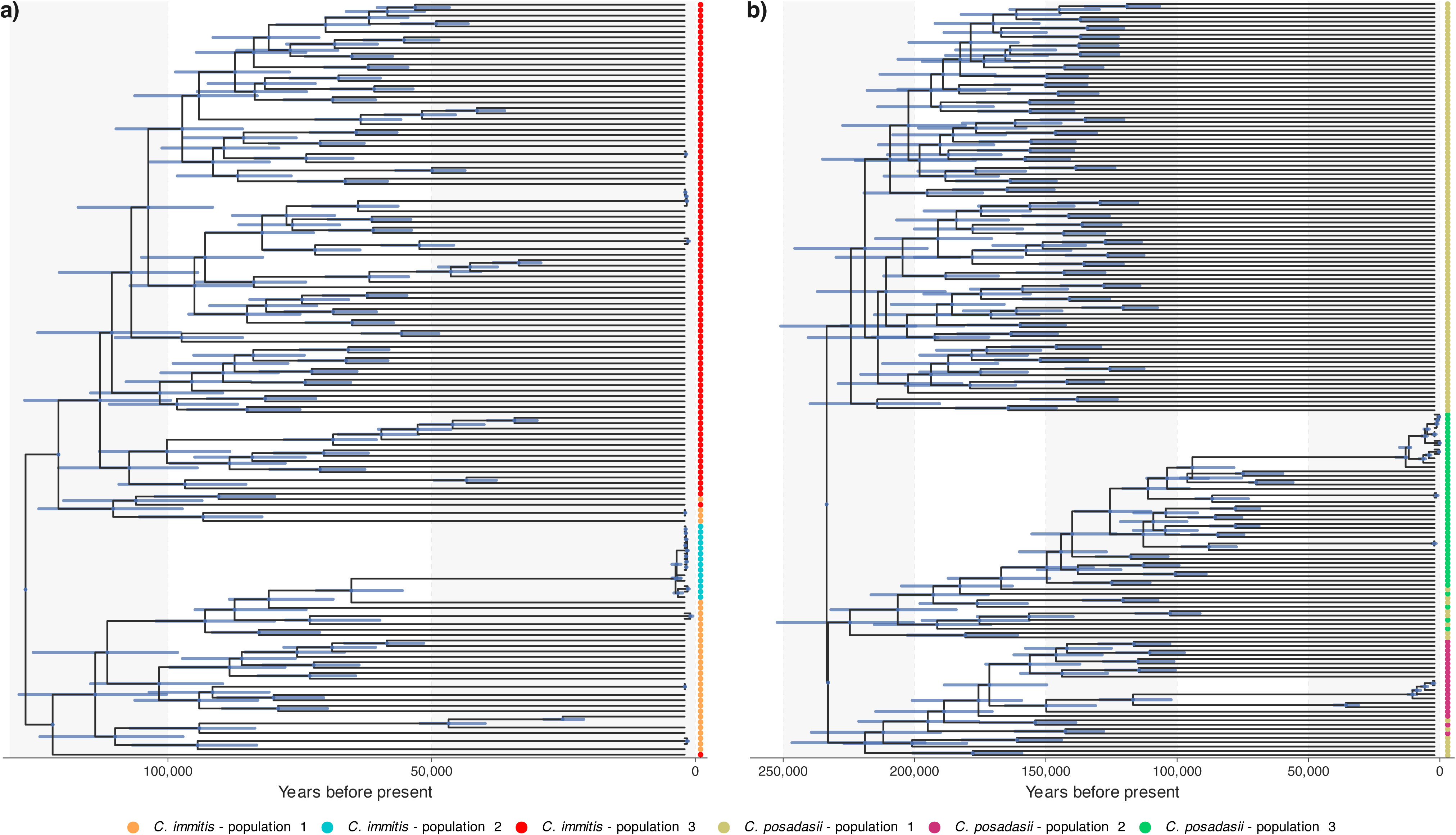
Time-scaled phylogenetic trees reconstructed from whole-genome SNP data for *C. immitis* (left) and *C. posadasii* (right). Branch lengths are scaled to years before present. Tip point color indicates population assignments inferred from population structure analyses. Purple bars at nodes indicate uncertainty (95% posterior density interval) in node age.

### Ancestral area reconstruction

Ancestral state reconstruction showed distinct geographic patterns of dispersal in both species (Fig. S3). In *C. immitis*, Populations 1 and 3 were inferred to originate in California with strong support (both p = 0.99), whereas Population 2 had a Pacific Northwest origin, with support split between Washington (p = 0.556) and Oregon (p = 0.444). Population 1 showed limited movement, with three inferred transitions from California to Washington, while Population 3 showed broader movement from California, mainly to Oregon and Washington.

Population 2 showed only local transitions between Oregon and Washington, consistent with circulation within the Pacific Northwest. In *C. posadasii*, Populations 1 and 2 were inferred to originate in Arizona with strong support (both p = 0.99), while Population 3 was inferred to originate in Texas (p = 0.98). Population 1 showed the broadest geographic spread, especially from Arizona to Oregon and Washington, whereas Population 3 moved mainly from Texas to Mexico.

### Mating Type Distribution Across Populations

Mating type was resolved for 308 of 312 isolates (98.7%) (Fig. 5). In *C. immitis*, populations 1 and 3 contained both mating types, with MAT1-1 frequencies of 46.9% (15/32) and 56.4% (53/94), respectively. In contrast, all 15 isolates in Population 2 had MAT1-2, consistent with clonality. In *C. posadasii*, all three populations contained both mating types, with MAT1-1 frequencies of 51.0% (53/104), 45.0% (9/20), and 55.8% (24/43) in Populations 1, 2, and 3, respectively. All 13 Brazilian isolates in Population 3 carried MAT1-1, and all five Venezuelan isolates in Population 2 carried MAT1-2, consistent with separate localized clonal expansions.

**Figure 5.**
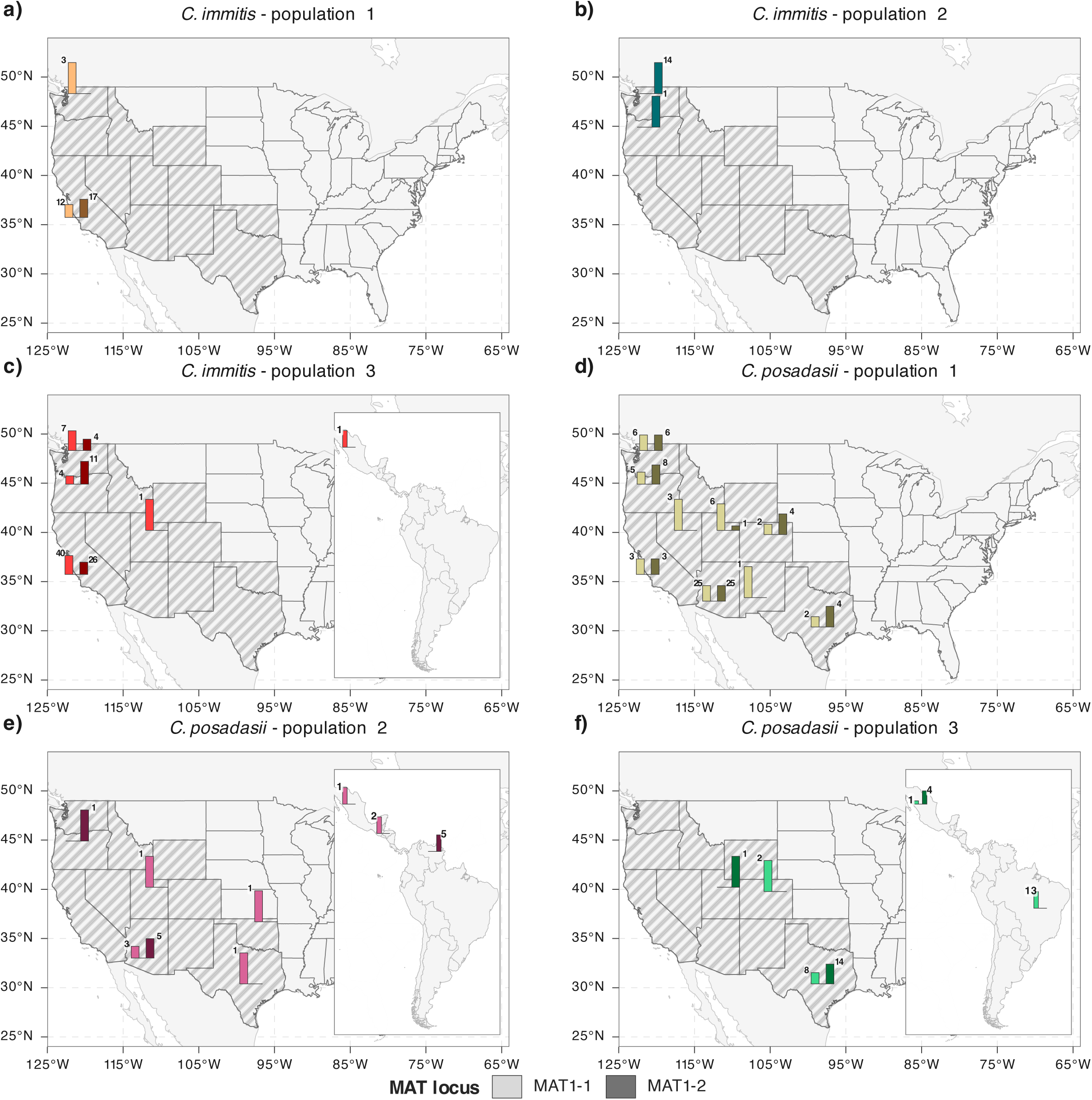
Barplots showing counts of MAT1-1 and MAT1-2 idiomorphs across four populations in (a) *C. immitis* and (b) *C. posadasii*.

### Historical demography

Historical effective population size increased across all *Coccidioides* populations, with distinct trajectories (Fig. 6). In *C. immitis*, Population 1 expanded from approximately 17,000 individuals 30,000 years ago to 160,000 by 10,000 years ago and then remained relatively stable. Population 2 showed a more recent expansion, from approximately 340 individuals 1,700 years ago to 8,400 at present. Population 3 showed the strongest expansion, increasing from approximately 14,000 individuals 30,000 years ago to 74,000 by 10,000 years ago, followed by a sharp rise to approximately 1.3 million at present. In *C. posadasii*, all three populations also expanded. Population 1 increased from approximately 120,000 individuals 30,000 years ago to 310,000 by 10,000 years ago and to 1.6 million near the present. Population 2 remained relatively stable until after 15,000 years ago before increasing to approximately 180,000 at present, while Population 3 increased more gradually to approximately 270,000 at present.

**Figure 6.**
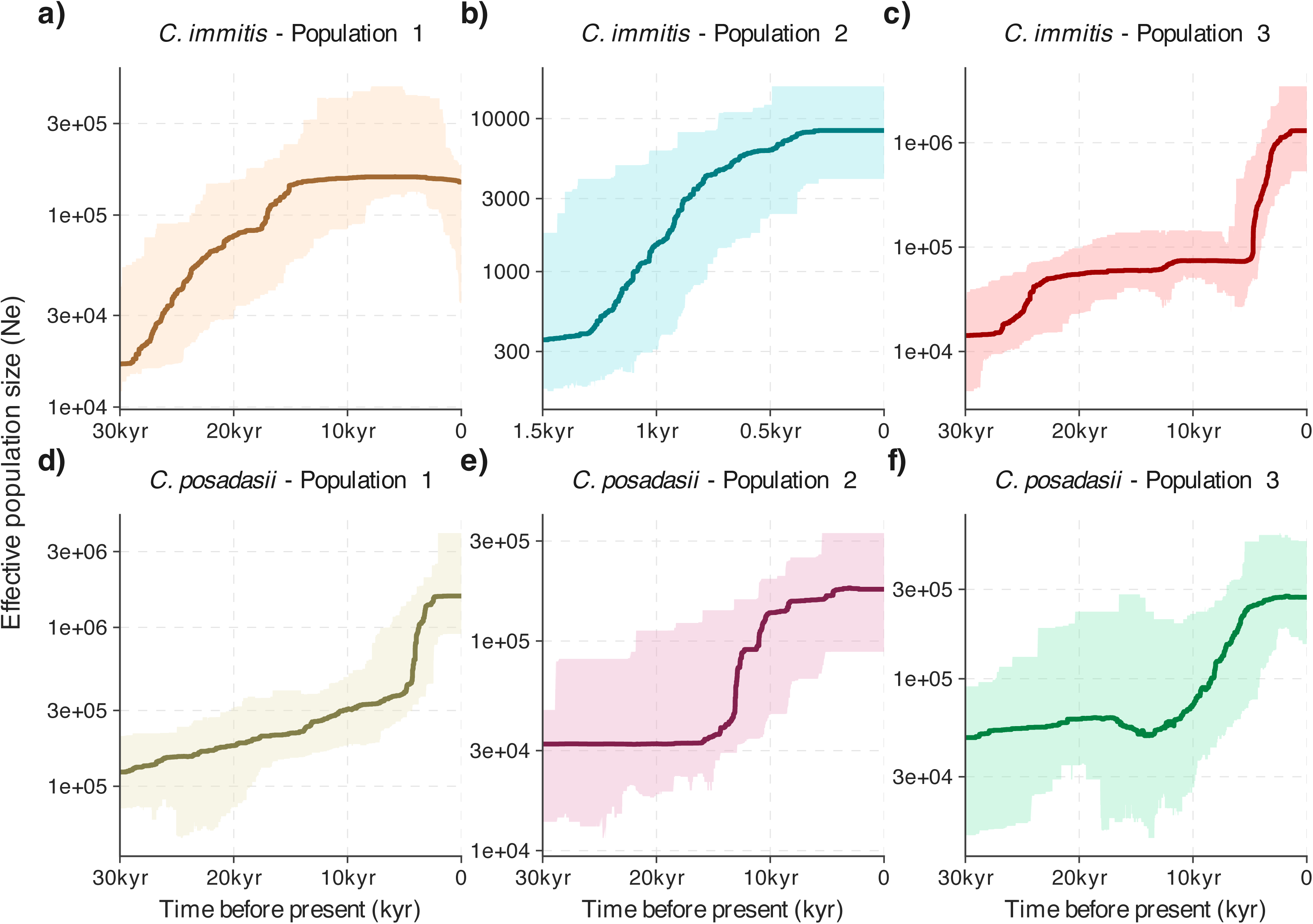
Changes in effective population size (Ne) through time estimated for each population using coalescent-based demographic inference. Solid lines represent median estimates of effective population size, and shaded regions indicate confidence intervals. Time is shown in thousands of years before present (kyr).

## Discussion

We show that *Coccidioides* populations are geographically structured but dynamic, with regionally associated populations, evidence of inter-regional gene flow, recent demographic expansion, and contrasting reproductive patterns across lineages. Together, these findings suggest that long-standing population differentiation coexists with ongoing genetic exchange and population growth, providing a genomic framework for understanding changing patterns of Valley fever endemicity.

In *C. immitis*, we recovered three major populations, including the recently emerged, clonal Pacific Northwest population, for which genomic and environmental data support locally-acquired infections from an environmental reservoir^4,20,54^. The finding that this population traces back to a common ancestor approximately 3,700 years ago, supports recent regional emergence rather than long-term endemicity^16^. In contrast, the other *C. immitis* populations showed broader geographic distributions and contained both mating types, consistent with the potential for sexual reproduction and recombination^21^. In *C. posadasii*, all three populations also contained both mating types, but geographically restricted sublineages showed evidence of clonality: all sampled Brazilian isolates within Population 3 carried MAT1-1^55,56^, and all sampled Venezuelan isolates within Population 2 carried MAT1-2,^22^ consistent with previous reports of reduced diversity in these groups. Two Guatemalan isolates within Population 2 also carried MAT1-1, raising the possibility of another geographically restricted clonal lineage, although broader sampling is needed to confirm this pattern.

We observed consistent evidence of gene flow between endemic regions, with dispersal largely radiating from established foci, including from California for *C. immitis* and Arizona and Texas for *C. posadasii*. Because sampling location reflects specimen submission rather than confirmed exposure location, these inferred transitions should be interpreted as evidence of genetic connectivity among regions rather than precise maps of infection origin^57,58^. Travel-associated infection probably contributed to some patterns in our clinical isolate collection, particularly for isolates submitted from lower-incidence regions^17,58,59^. For example, one Utah *C. immitis* isolate from our study was likely a travel-acquired infection, from an individual reporting recent travel to San Diego, where *C. immitis* is endemic, consistent with previous reports^18^.

However, travel alone may not explain all observed structure: repeated inferred transitions from California into Oregon and Washington in *C. immitis*, and from Arizona or Texas into multiple western and international localities in *C. posadasii*, suggest that broader dispersal processes may contribute to these patterns. The Venezuela and Brazil isolates were assigned to different *C. posadasii* populations and carried different mating types, consistent with multiple long-distance dispersal histories rather than a single *C. posadasii* introduction into South America. More broadly, fungal dispersal may occur through multiple hypothesized mechanisms, including localized aerosolization from disturbed soil^60,61^ and possible animal-associated processes^61,62^, although the specific mechanisms underlying long-distance movement remain unresolved.

A central finding of this study is that demographic expansion was detected across all major *Coccidioides* populations, although the timing and magnitude of growth differed among lineages. These trends broadly overlap with post–Last Glacial Maximum climatic changes in North America, including loss of glaciers and warming with regionally variable increases in effective moisture during the transition into the Holocene^63,64^. The southwestern United States was also wetter during parts of this period and supported abundant large mammals, many of which disappeared during the late Pleistocene^65,66^. Because *Coccidioides* can associate with mammalian hosts, these ecological disruptions may have altered host availability. Together, changes in climate, vegetation, soil conditions, and mammalian communities may have increased habitat suitability and connectivity for *Coccidioides*. Although deeper-time demographic inference is limited by coalescent resolution, the observed patterns are consistent with environmentally driven expansion of fungal populations. These findings raise the possibility that ongoing climate change could further alter the geographic distribution and population dynamics of *Coccidioides*^9,67,68^.

Time-calibrated phylogenies estimated that sampled *C. posadasii* lineages shared a substantially older common ancestor than sampled *C. immitis* lineages. Divergence-time estimates were sensitive to LD pruning, with analyses retaining linked sites producing older estimates that approached previous reports. This pattern suggests that linkage can influence branch-length scaling and divergence-time inference, highlighting the importance of model assumptions when interpreting evolutionary timescales^69–71^.

These findings point to priorities for future research. Paired clinical and environmental sampling will be essential to resolve exposure sources, identify environmental reservoirs, and distinguish locally acquired from travel-associated infections. Improved estimates of generation time and mutation rate will also be needed to calibrate evolutionary timescales more precisely, because the effective generation time of *Coccidioides* in nature remains uncertain^72,7361^. Finally, ecological studies linking genomic evidence of population growth to environmental suitability will be critical for anticipating future changes in *Coccidioides* distribution and human infection risk.

By showing that *Coccidioides* populations are both geographically structured and demographically expanding, this study provides a genomic framework for interpreting changing Valley fever risk. Future surveillance that links clinical genomes, environmental sampling, and patient exposure data will be essential for distinguishing local emergence from travel-associated disease and for detecting shifts in endemicity.

## Contributors

All authors conceived and designed the study. MH, KH, and KSW collected the data. EMF and KSW developed the methodology and EMF performed the analyses. KH, MH, and KSW supervised clinical data generation. All authors contributed to data interpretation. EMF and KSW wrote the initial draft of the manuscript. All authors reviewed, revised, and approved the final version. EMF and KSW had full access to all the data and verified the underlying data.

## Data sharing

Raw sequencing data are available in the Sequence Read Archive (SRA) under BioProject accession PRJNA1459073. Accession numbers for sequences generated in this study range from SAMN57541207 to SAMN57541421 and are listed in Table S1.

## Research in Context

### Evidence before this study

We searched PubMed, Web of Science, and Google Scholar from Jan 1, 2000, to Mar 20, 2026, using the terms “*Coccidioides*”, “coccidioidomycosis”, “phylogeography”, and “genomics”, without language restrictions. Previous genomic studies have provided important insights into population structure, evolutionary history, and dispersal, including evidence of movement into non-endemic regions. Together, these studies provide a foundation for integrating genetic structure, geographic movement, demographic history, and reproductive patterns across clinically relevant populations of *C. immitis* and *C. posadasii*.

### Added value of this study

We combined genomes from isolates collected via prospective national sampling with publicly available genomes to generate a unified dataset for *C. immitis* and *C. posadasii*. We used this dataset to examine population structure, geographic movement, reproductive patterns, and recent expansion. major populations were genetically distinct but connected by shared ancestry and/or gene flow, with movement largely originating from high-incidence regions. We also found evidence of recent demographic expansion across populations and localized clonal spread in specific populations.

### Implications of all the available evidence

The available evidence suggests that geographically structured *Coccidioides* populations, including both recombining and clonal populations, are undergoing recent demographic expansion. Genomic surveillance will be critical to track shifts in distribution and anticipate changes in disease risk under evolving environmental conditions.

## Supporting information

Supplementary table of genome accession numbers, species, geographic origins, and sources, with additional phylogenetic and ancestral-area analyses.

## Data Availability

All raw sequencing data generated in this study are available in the NCBI Sequence Read Archive under BioProject accession PRJNA1459073. Accession numbers for the newly generated sequences are provided in Table S1. All publicly available genomes included in the analyses are also listed in Table S1 with their corresponding accession numbers.

## References

1 Johnson RH, Sharma R, Kuran R, Fong I, Heidari A. Coccidioidomycosis: A review. Journal of Investigative Medicine. 2021; 69: 316–23.

2 Crum NF. Coccidioidomycosis: A Contemporary Review. Infect. Dis. Ther. 2022; 11: 713–42.

3 Centers for Disease Control and Prevention. Reported cases of Valley fever (coccidioidomycosis). U.S. Department of Health and Human Services. 2024. https://www.cdc.gov/valley-fever/php/statistics/index.html (accessed Dec 20, 2025).

4 Marsden-Haug N, Goldoft M, Ralston C, et al. Coccidioidomycosis acquired in Washington State. Clinical Infectious Diseases 2013; 56: 847–50.

5 Johnson SM, Carlson EL, Fisher FS, Pappagianis D. Demonstration of Coccidioides immitis and Coccidioides posadasii DNA in soil samples collected from Dinosaur National Monument, Utah. Med Mycol 2014; 52: 610–7.

6 Ampel NM. What’s Behind the Increasing Rates of Coccidioidomycosis in Arizona and California? Curr Infect Dis Rep 2010; 12: 211–6.

7 Williams SL, Benedict K, Jackson BR, et al. Estimated Burden of Coccidioidomycosis. JAMA Netw Open 2025; 8. DOI:10.1001/jamanetworkopen.2025.13572.

8 Benedict K, McCotter OZ, Brady S, et al. Surveillance for Coccidioidomycosis — United States, 2011–2017. MMWR Surveillance Summaries 2019; 68: 1–15.

9 Gorris ME, Treseder KK, Zender CS, Randerson JT. Expansion of Coccidioidomycosis Endemic Regions in the United States in Response to Climate Change. Geohealth 2019; 3: 308–27.

10 Lee PS, Swain DL, Johnson R. Climate Change and Coccidioidomycosis. JAMA. 2025; 333: 997–8.

11 Weaver E, Kolivras KN, Thomas RQ, Thomas VA, Abbas KM. Environmental factors affecting ecological niche of Coccidioides species and spatial dynamics of valley fever in the United States. Spat Spatiotemporal Epidemiol 2020; 32: 100317.

12 Head JR, Sondermeyer-Cooksey G, Heaney AK, et al. Effects of precipitation, heat, and drought on incidence and expansion of coccidioidomycosis in western USA: a longitudinal surveillance study. Lancet Planet Health 2022; 6: e793–803.

13 Gorris ME, Ardon-Dryer K, Campuzano A, et al. Advocating for Coccidioidomycosis to Be a Reportable Disease Nationwide in the United States and Encouraging Disease Surveillance across North and South America. Journal of Fungi 2023; 9. DOI:10.3390/jof9010083.

14 Hossain T, Ibarra-Mejia G, Romero-Olivares AL, Gill TE. Environmental Detection of Coccidioides: Challenges and Opportunities. Environments-MDPI. 2025; 12. DOI:10.3390/environments12080258.

15 Dobos RR, Benedict K, Jackson BR, McCotter OZ. Using soil survey data to model potential Coccidioides soil habitat and inform Valley fever epidemiology. PLoS One 2021; 16. DOI:10.1371/journal.pone.0247263.

16 Oltean HN, Etienne KA, Roe CC, et al. Utility of whole-genome sequencing to ascertain locally acquired cases of coccidioidomycosis, Washington, USA. Emerg Infect Dis 2019; 25: 501–6.

17 Engelthaler DM, Roe CC, Hepp CM, et al. Local population structure and patterns of Western Hemisphere dispersal for Coccidioides spp., the fungal cause of valley fever. mBio 2016; 7. DOI:10.1128/mBio.00550-16.

18 Fonseca EM, Fox S, Carey AL, et al. Coccidioides genomes from low-incidence states reveal complex migration history across the western United States. Microbiol Spectr 2025; published online Dec 2. DOI:10.1128/spectrum.01822-25.

19 Fisher MC, Koenig GL, White TJ, et al. Biogeographic range expansion into South America by *Coccidioides immitis* mirrors New World patterns of human migration. Proceedings of the National Academy of Sciences 2001; 98: 4558–62.

20 Litvintseva AP, Marsden-Haug N, Hurst S, et al. Valley Fever: Finding New Places for an Old Disease: Coccidioides immitis Found in Washington State Soil Associated With Recent Human Infection. Clinical Infectious Diseases 2015; 60: e1–3.

21 Mandel MA, Barker BM, Kroken S, Rounsley SD, Orbach MJ. Genomic and Population Analyses of the Mating Type Loci in *Coccidioides* Species Reveal Evidence for Sexual Reproduction and Gene Acquisition. Eukaryot Cell 2007; 6: 1189–99.

22 Teixeira MM, Alvarado P, Roe CC, et al. Population Structure and Genetic Diversity among Isolates of *Coccidioides posadasii* in Venezuela and Surrounding Regions. mBio 2019; 10. DOI:10.1128/mBio.01976-19.

23 Wengenack NL., Lainhart William, Wiederhold NP. Principles and procedures for detection and culture of fungi in clinical specimens. Clinical and Lboratory Standards Institute, 2021.

24 Marchetti M, Fonseca EM, Hanson KE, Barker B, Walter KS. The impact of bioinformatic choices on Coccidioides variant identification accuracy. Microbiol Spectr 2025; 13. DOI:10.1128/spectrum.01232-25.

25 Wood DE, Lu J, Langmead B. Improved metagenomic analysis with Kraken 2. Genome Biol 2019; 20. DOI:10.1186/s13059-019-1891-0.

26 Sharpton TJ, Stajich JE, Rounsley SD, et al. Comparative genomic analyses of the human fungal pathogens Coccidioides and their relatives. Genome Res 2009; 19: 1722–31.

27 Neafsey DE, Barker BM, Sharpton TJ, et al. Population genomic sequencing of Coccidioides fungi reveals recent hybridization and transposon control. Genome Res 2010; 20: 938–46.

28 de Melo Teixeira M, Lang BF, Matute DR, Stajich JE, Barker BM. Mitochondrial genomes of the human pathogens *Coccidioides immitis* and *Coccidioides posadasii*. G3 2021; 11. DOI:10.1093/g3journal/jkab132.

29 Melo Teixeira M, Stajich JE, Sahl JW, et al. A chromosomal-level reference genome of the widely utilized Coccidioides posadasii laboratory strain ‘Silveira’. G3: Genes, Genomes, Genetics 2022; 12. DOI:10.1093/g3journal/jkac031.

30 Li H, Durbin R. Fast and accurate short read alignment with Burrows-Wheeler transform. Bioinformatics 2009; 25: 1754–60.

31 McKenna A, Hanna M, Banks E, et al. The genome analysis toolkit: A MapReduce framework for analyzing next-generation DNA sequencing data. Genome Res 2010; 20: 1297–303.

32 Kurtz S, Phillippy A, Delcher AL, et al. Open Access Versatile and open software for comparing large genomes. 2004 http://www.tigr.org/software/mummer.

33 Cingolani P, Platts A, Wang LL, et al. A program for annotating and predicting the effects of single nucleotide polymorphisms, SnpEff: SNPs in the genome of Drosophila melanogaster strain w1118; iso-2; iso-3. Fly (Austin) 2012; 6: 80–92.

34 Chang CC, Chow CC, Tellier LC, Vattikuti S, Purcell SM, Lee JJ. Second-generation PLINK: rising to the challenge of larger and richer datasets. Gigascience 2015; 4. DOI:10.1186/s13742-015-0047-8.

35 Frichot E, François O. LEA: An R package for landscape and ecological association studies. Methods Ecol Evol 2015; 6: 925–9.

36 Frichot E, Mathieu F, Trouillon T, Bouchard G, François O. Fast and efficient estimation of individual ancestry coefficients. Genetics 2014; 196: 973–83.

37 Jombart T. adegenet: a R package for the multivariate analysis of genetic markers. Bioinformatics 2008; 24: 1403–5.

38 Carstens BC, Pelletier TA, Reid NM, Satler JD. How to fail at species delimitation. Mol Ecol 2013; 22: 4369–83.

39 Minh BQ, Schmidt HA, Chernomor O, et al. IQ-TREE 2: New Models and Efficient Methods for Phylogenetic Inference in the Genomic Era. Mol Biol Evol 2020; 37: 1530–4.

40 Kalyaanamoorthy S, Minh BQ, Wong TKF, von Haeseler A, Jermiin LS. ModelFinder: fast model selection for accurate phylogenetic estimates. Nat Methods 2017; 14: 587–9.

41 Hoang DT, Chernomor O, von Haeseler A, Minh BQ, Vinh LS. UFBoot2: Improving the Ultrafast Bootstrap Approximation. Mol Biol Evol 2018; 35: 518–22.

42 Sagulenko P, Puller V, Neher RA. TreeTime: Maximum-likelihood phylodynamic analysis. Virus Evol 2018; 4. DOI:10.1093/ve/vex042.

43 Paradis E. Pegas: An R package for population genetics with an integrated-modular approach. Bioinformatics 2010; 26: 419–20.

44 Wang Y-T, Wu H-X, Chomnunti P. The role of ancestral state reconstruction in mycological researches: methods and insights. Stud Fungi 2025; 10: e022.

45 Sheikh S, Khan FK, Bahram M, Ryberg M. Impact of model assumptions on the inference of the evolution of ectomycorrhizal symbiosis in fungi. Sci Rep 2022; 12: 22043.

46 Barber AE, Sae-Ong T, Kang K, et al. Aspergillus fumigatus pan-genome analysis identifies genetic variants associated with human infection. Nat Microbiol 2021; 6: 1526–36.

47 Vasimuddin Md, Misra S, Li H, Aluru S. Efficient Architecture-Aware Acceleration of BWA-MEM for Multicore Systems. In: 2019 IEEE International Parallel and Distributed Processing Symposium (IPDPS). IEEE, 2019: 314–24.

48 Danecek P, Bonfield JK, Liddle J, et al. Twelve years of SAMtools and BCFtools. Gigascience 2021; 10. DOI:10.1093/gigascience/giab008.

49 Li H, Handsaker B, Wysoker A, et al. The Sequence Alignment/Map format and SAMtools. Bioinformatics 2009; 25: 2078–9.

50 Liu X, Fu YX. Stairway Plot 2: demographic history inference with folded SNP frequency spectra. Genome Biol 2020; 21: 1–9.

51 Kruger SE, Ruberto I, Williamson T, Remais J V., Heaney AK, Head JR. Regional Increases in Incidence of Coccidioidomycosis (Valley Fever) — Arizona, 2005–2022. MMWR Morb Mortal Wkly Rep 2026; 75: 85–91.

52 Galgiani JN, Ampel NM, Blair JE, et al. Coccidioidomycosis. 2005 https://academic.oup.com/cid/article/41/9/1217/277222.

53 Teixeira MM, Barker BM. Use of population genetics to assess the ecology, evolution, and population structure of Coccidioides. Emerg Infect Dis 2016; 22: 1022–30.

54 McCotter OZ, Benedict K, Engelthaler DM, et al. Update on the Epidemiology of coccidioidomycosis in the United States. Med. Mycol. 2019; 57: S30–40.

55 Eulálio KD, Kollath DR, Martins LMS, et al. Epidemiological, clinical, and genomic landscape of coccidioidomycosis in northeastern Brazil. Nat Commun 2024; 15. DOI:10.1038/s41467-024-47388-0.

56 Fraser HB. Gene expression drives local adaptation in humans. Genome Res 2013; 23: 1089–96.

57 Barker BM, Rajan S, De Melo Teixeira M, et al. Coccidioidal Meningitis in New York Traced to Texas by Fungal Genomic Analysis. Clinical Infectious Diseases 2019; 69: 1060–2.

58 Monroy-Nieto J, Gade L, Benedict K, et al. Genomic Epidemiology Linking Nonendemic Coccidioidomycosis to Travel. Emerg Infect Dis 2023; 29: 110–7.

59 Qazi I, Hancock N. Disseminated coccidioidomycosis following travel to an endemic region and COVID-19 infection: Case report and case-based literature review. Medical Reports 2025; 9: 100164.

60 Porter WT, Gade L, Montfort P, et al. Understanding the exposure risk of aerosolized Coccidioides in a Valley fever endemic metropolis. Sci Rep 2024; 14. DOI:10.1038/s41598-024-51407-x.

61 Kollath DR, Miller KJ, Barker BM. The mysterious desert dwellers: Coccidioides immitis and Coccidioides posadasii, causative fungal agents of coccidioidomycosis. Virulence. 2019; 10: 222–33.

62 Taylor JW, Barker BM. The endozoan, small-mammal reservoir hypothesis and the life cycle of Coccidioides species. Med Mycol 2019; 57: S16–20.

63 Clark PU, Shakun JD, Baker PA, et al. Global climate evolution during the last deglaciation. Proceedings of the National Academy of Sciences 2012; 109. DOI:10.1073/pnas.1116619109.

64 Clark PU, Dyke AS, Shakun JD, et al. The Last Glacial Maximum. Science (1979) 2009; 325: 710–4.

65 Grayson DK. The Late Quaternary biogeographic histories of some Great Basin mammals (western USA). Quat Sci Rev 2006; 25: 2964–91.

66 Stewart M, Carleton WC, Groucutt HS. Climate change, not human population growth, correlates with Late Quaternary megafauna declines in North America. Nat Commun 2021; 12: 965.

67 Head JR, Sondermeyer-Cooksey G, Heaney AK, et al. Effects of precipitation, heat, and drought on incidence and expansion of coccidioidomycosis in western USA: a longitudinal surveillance study. Lancet Planet Health 2022; 6: e793–803.

68 Gorris ME, Cat LA, Zender CS, Treseder KK, Randerson JT. Coccidioidomycosis Dynamics in Relation to Climate in the Southwestern United States. Geohealth 2018; 2: 6–24.

69 Slatkin M. Linkage disequilibrium — understanding the evolutionary past and mapping the medical future. Nat Rev Genet 2008; 9: 477–85.

70 Zou Y, Zhang Z, Zeng Y, et al. Common Methods for Phylogenetic Tree Construction and Their Implementation in R. Bioengineering. 2024; 11. DOI:10.3390/bioengineering11050480.

71 Magee AF, Hilton SK, Dewitt WS. Robustness of Phylogenetic Inference to Model Misspecification Caused by Pairwise Epistasis. Mol Biol Evol 2021; 38: 4603–15.

72 Mead HL, Van Dyke MCC, Barker BM. Proper Care and Feeding of *Coccidioides*: A Laboratorian’s Guide to Cultivating the Dimorphic Stages of *C. immitis* and *C. posadasii*. Curr Protoc Microbiol 2020; 58. DOI:10.1002/cpmc.113.

73 Higgins Keppler EA, Mead HL, Barker BM, Bean HD. Life Cycle Dominates the Volatilome Character of Dimorphic Fungus Coccidioides spp. mSphere 2021; 6. DOI:10.1128/msphere.00040-21.

