## Supplementary table of genome accession numbers, species, geographic origins, and sources, with additional phylogenetic and ancestral-area analyses. for "Genomic insights into the population structure and recent expansion of *Coccidioides* in the United States"

### **Table S1.** Accession numbers, species identification, and geographic origin for all *Coccidioides* genomes used in this study. Accession numbers correspond to entries in the NCBI Sequence Read Archive (SRA). Species designations (*Coccidioides immitis* or *Coccidioides posadasii*) were inferred using the cocci-call pipeline. Geographic origin refers to the state or country where the isolate was originally collected.

| **Accession Number** | **Species** | **Geographic Origin** | **Genome Source** |
| --- | --- | --- | --- |
| SAMN52167826 | *Coccidioides immitis* | California | This Study |
| SAMN57541207 | *Coccidioides immitis* | California | This Study |
| SAMN52167869 | *Coccidioides immitis* | Utah | This Study |
| SAMN57541209 | *Coccidioides immitis* | California | This Study |
| SAMN57541210 | *Coccidioides immitis* | California | This Study |
| SRR1292227 | *Coccidioides immitis* | Washington | Previously published |
| SRR21204704 | *Coccidioides immitis* | Oregon | Previously published |
| SRR1292225 | *Coccidioides immitis* | Washington | Previously published |
| SRR1292219 | *Coccidioides immitis* | California | Previously published |
| SRR8530937 | *Coccidioides immitis* | Washington | Previously published |
| SRR21204723 | *Coccidioides immitis* | Oregon | Previously published |
| SRR21204674 | *Coccidioides immitis* | Washington | Previously published |
| SRR21204691 | *Coccidioides immitis* | Washington | Previously published |
| SRR1292224 | *Coccidioides immitis* | Washington | Previously published |
| SAMN57541213 | *Coccidioides immitis* | California | This Study |
| SAMN57541214 | *Coccidioides immitis* | California | This Study |
| SAMN52167837 | *Coccidioides immitis* | California | This Study |
| SAMN57541215 | *Coccidioides immitis* | California | This Study |
| SAMN57541216 | *Coccidioides immitis* | California | This Study |
| SAMN57541217 | *Coccidioides immitis* | California | This Study |
| SAMN57541219 | *Coccidioides immitis* | California | This Study |
| SAMN57541220 | *Coccidioides immitis* | California | This Study |
| SAMN52167827 | *Coccidioides immitis* | California | This Study |
| SAMN57541221 | *Coccidioides immitis* | California | This Study |
| SRR21204693 | *Coccidioides immitis* | Oregon | Previously published |
| SAMN57541222 | *Coccidioides immitis* | California | This Study |
| SAMN57541224 | *Coccidioides immitis* | California | This Study |
| SAMN57541226 | *Coccidioides immitis* | California | This Study |
| SAMN57541227 | *Coccidioides immitis* | California | This Study |
| SAMN52167835 | *Coccidioides immitis* | California | This Study |
| SAMN57541229 | *Coccidioides immitis* | California | This Study |
| SAMN57541230 | *Coccidioides immitis* | California | This Study |
| SAMN57541231 | *Coccidioides immitis* | California | This Study |
| SRR21204698 | *Coccidioides immitis* | Washington | Previously published |
| SRR3468019 | *Coccidioides immitis* | California | Previously published |
| SRR21204680 | *Coccidioides immitis* | Oregon | Previously published |
| SRR7206605 | *Coccidioides immitis* | Washington | Previously published |
| SRR3468081 | *Coccidioides immitis* | California | Previously published |
| SRR8530933 | *Coccidioides immitis* | Washington | Previously published |
| SRR21204677 | *Coccidioides immitis* | Washington | Previously published |
| SRR21295063 | *Coccidioides immitis* | Oregon | Previously published |
| SRR8530936 | *Coccidioides immitis* | Washington | Previously published |
| SRR21204679 | *Coccidioides immitis* | Oregon | Previously published |
| SRR21204687 | *Coccidioides immitis* | Oregon | Previously published |
| SRR3468071 | *Coccidioides immitis* | California | Previously published |
| SRR3468049 | *Coccidioides immitis* | California | Previously published |
| SRR1292218 | *Coccidioides immitis* | California | Previously published |
| SRR7206603 | *Coccidioides immitis* | Washington | Previously published |
| SRR21204713 | *Coccidioides immitis* | Oregon | Previously published |
| SRR21204709 | *Coccidioides immitis* | Washington | Previously published |
| SRR21204705 | *Coccidioides immitis* | Oregon | Previously published |
| SRR21204694 | *Coccidioides immitis* | Oregon | Previously published |
| SRR21204695 | *Coccidioides immitis* | Washington | Previously published |
| SRR8530934 | *Coccidioides immitis* | Washington | Previously published |
| SRR21204675 | *Coccidioides immitis* | Washington | Previously published |
| SRR1292228 | *Coccidioides immitis* | Washington | Previously published |
| SRR3468021 | *Coccidioides immitis* | Mexico | Previously published |
| SRR8530935 | *Coccidioides immitis* | Washington | Previously published |
| SRR21204689 | *Coccidioides immitis* | Washington | Previously published |
| SRR21204701 | *Coccidioides immitis* | Oregon | Previously published |
| SRR21204683 | *Coccidioides immitis* | Oregon | Previously published |
| SRR7206599 | *Coccidioides immitis* | Washington | Previously published |
| SRR21204692 | *Coccidioides immitis* | Oregon | Previously published |
| SRR21204720 | *Coccidioides immitis* | Washington | Previously published |
| SRR3468038 | *Coccidioides immitis* | California | Previously published |
| SRR21204700 | *Coccidioides immitis* | Washington | Previously published |
| SRR1292226 | *Coccidioides immitis* | Washington | Previously published |
| SRR7206601 | *Coccidioides immitis* | Washington | Previously published |
| SRR21204703 | *Coccidioides immitis* | Oregon | Previously published |
| SRR7206598 | *Coccidioides immitis* | Washington | Previously published |
| SRR21204699 | *Coccidioides immitis* | Oregon | Previously published |
| SRR21204731 | *Coccidioides immitis* | Washington | Previously published |
| SRR3468016 | *Coccidioides immitis* | California | Previously published |
| SAMN57541233 | *Coccidioides immitis* | California | This Study |
| SAMN57541234 | *Coccidioides immitis* | California | This Study |
| SAMN57541235 | *Coccidioides immitis* | California | This Study |
| SAMN57541236 | *Coccidioides immitis* | California | This Study |
| SAMN57541237 | *Coccidioides immitis* | California | This Study |
| SAMN52167842 | *Coccidioides immitis* | California | This Study |
| SAMN57541239 | *Coccidioides immitis* | California | This Study |
| SAMN57541240 | *Coccidioides immitis* | California | This Study |
| SAMN57541241 | *Coccidioides immitis* | California | This Study |
| SAMN52167839 | *Coccidioides immitis* | California | This Study |
| SAMN57541242 | *Coccidioides immitis* | California | This Study |
| SAMN57541243 | *Coccidioides immitis* | California | This Study |
| SAMN57541244 | *Coccidioides immitis* | California | This Study |
| SAMN57541245 | *Coccidioides immitis* | California | This Study |
| SAMN52167840 | *Coccidioides immitis* | California | This Study |
| SAMN57541246 | *Coccidioides immitis* | California | This Study |
| SAMN52167841 | *Coccidioides immitis* | California | This Study |
| SAMN57541247 | *Coccidioides immitis* | California | This Study |
| SAMN57541249 | *Coccidioides immitis* | California | This Study |
| SAMN57541250 | *Coccidioides immitis* | California | This Study |
| SAMN57541251 | *Coccidioides immitis* | California | This Study |
| SAMN52167847 | *Coccidioides immitis* | California | This Study |
| SAMN57541253 | *Coccidioides immitis* | California | This Study |
| SAMN57541254 | *Coccidioides immitis* | California | This Study |
| SAMN57541255 | *Coccidioides immitis* | California | This Study |
| SAMN57541258 | *Coccidioides immitis* | California | This Study |
| SAMN57541259 | *Coccidioides immitis* | California | This Study |
| SAMN57541261 | *Coccidioides immitis* | California | This Study |
| SAMN57541262 | *Coccidioides immitis* | Oregon | This Study |
| SAMN57541263 | *Coccidioides immitis* | California | This Study |
| SAMN57541264 | *Coccidioides immitis* | California | This Study |
| SAMN57541266 | *Coccidioides immitis* | California | This Study |
| SAMN52167843 | *Coccidioides immitis* | California | This Study |
| SAMN57541267 | *Coccidioides immitis* | California | This Study |
| SAMN52167829 | *Coccidioides immitis* | California | This Study |
| SAMN52167838 | *Coccidioides immitis* | California | This Study |
| SAMN57541269 | *Coccidioides immitis* | California | This Study |
| SAMN52167832 | *Coccidioides immitis* | California | This Study |
| SAMN57541270 | *Coccidioides immitis* | Washington | This Study |
| SAMN57541271 | *Coccidioides immitis* | California | This Study |
| SAMN57541272 | *Coccidioides immitis* | California | This Study |
| SAMN57541273 | *Coccidioides immitis* | California | This Study |
| SAMN57541274 | *Coccidioides immitis* | California | This Study |
| SAMN57541275 | *Coccidioides immitis* | California | This Study |
| SAMN57541277 | *Coccidioides immitis* | California | This Study |
| SAMN57541278 | *Coccidioides immitis* | California | This Study |
| SAMN52167828 | *Coccidioides immitis* | California | This Study |
| SAMN57541279 | *Coccidioides immitis* | California | This Study |
| SAMN57541280 | *Coccidioides immitis* | California | This Study |
| SAMN52167845 | *Coccidioides immitis* | California | This Study |
| SAMN57541281 | *Coccidioides immitis* | California | This Study |
| SAMN57541282 | *Coccidioides immitis* | California | This Study |
| SAMN57541283 | *Coccidioides immitis* | California | This Study |
| SAMN57541284 | *Coccidioides immitis* | California | This Study |
| SAMN57541286 | *Coccidioides immitis* | California | This Study |
| SAMN57541287 | *Coccidioides immitis* | California | This Study |
| SAMN57541289 | *Coccidioides immitis* | California | This Study |
| SAMN57541290 | *Coccidioides immitis* | California | This Study |
| SAMN57541291 | *Coccidioides immitis* | California | This Study |
| SAMN57541293 | *Coccidioides immitis* | California | This Study |
| SAMN57541294 | *Coccidioides immitis* | California | This Study |
| SAMN52167833 | *Coccidioides immitis* | California | This Study |
| SAMN57541295 | *Coccidioides immitis* | California | This Study |
| SAMN57541296 | *Coccidioides immitis* | California | This Study |
| SAMN57541298 | *Coccidioides immitis* | California | This Study |
| SAMN52167844 | *Coccidioides immitis* | California | This Study |
| SRR3468024 | *Coccidioides posadasii* | Arizona | Previously published |
| SRR3468043 | *Coccidioides posadasii* | Arizona | Previously published |
| SRR21204686 | *Coccidioides posadasii* | Washington | Previously published |
| SRR21204702 | *Coccidioides posadasii* | Oregon | Previously published |
| SRR21295062 | *Coccidioides posadasii* | Oregon | Previously published |
| SRR21204696 | *Coccidioides posadasii* | Oregon | Previously published |
| SAMN52167852 | *Coccidioides posadasii* | Colorado | This Study |
| SAMN52167853 | *Coccidioides posadasii* | Colorado | This Study |
| SRR21204714 | *Coccidioides posadasii* | Oregon | Previously published |
| SRR3468044 | *Coccidioides posadasii* | Arizona | Previously published |
| SAMN57541299 | *Coccidioides posadasii* | Arizona | This Study |
| SAMN57541300 | *Coccidioides posadasii* | Arizona | This Study |
| SRR21292482 | *Coccidioides posadasii* | Washington | Previously published |
| SRR3468064 | *Coccidioides posadasii* | Mexico | Previously published |
| SRR6830887 | *Coccidioides posadasii* | Mexico | Previously published |
| SRR3468028 | *Coccidioides posadasii* | Arizona | Previously published |
| SRR21292483 | *Coccidioides posadasii* | Washington | Previously published |
| SRR3468036 | *Coccidioides posadasii* | Arizona | Previously published |
| SRR21295066 | *Coccidioides posadasii* | Oregon | Previously published |
| SRR3468031 | *Coccidioides posadasii* | Arizona | Previously published |
| SRR3468041 | *Coccidioides posadasii* | Arizona | Previously published |
| SRR21204685 | *Coccidioides posadasii* | Oregon | Previously published |
| SRR21204706 | *Coccidioides posadasii* | Oregon | Previously published |
| SRR21204725 | *Coccidioides posadasii* | Oregon | Previously published |
| SRR3468056 | *Coccidioides posadasii* | Arizona | Previously published |
| SRR3468063 | *Coccidioides posadasii* | Arizona | Previously published |
| SRR3468023 | *Coccidioides posadasii* | Arizona | Previously published |
| SRR3468048 | *Coccidioides posadasii* | Texas | Previously published |
| SRR21204711 | *Coccidioides posadasii* | Washington | Previously published |
| SRR3468030 | *Coccidioides posadasii* | Arizona | Previously published |
| SRR3468066 | *Coccidioides posadasii* | Mexico | Previously published |
| SRR3468045 | *Coccidioides posadasii* | Arizona | Previously published |
| SRR21204681 | *Coccidioides posadasii* | Washington | Previously published |
| SRR3468069 | *Coccidioides posadasii* | Texas | Previously published |
| SRR3468035 | *Coccidioides posadasii* | Arizona | Previously published |
| SRR3468029 | *Coccidioides posadasii* | Arizona | Previously published |
| SRR21292480 | *Coccidioides posadasii* | Washington | Previously published |
| SRR21204710 | *Coccidioides posadasii* | Washington | Previously published |
| SRR3468075 | *Coccidioides posadasii* | Texas | Previously published |
| SRR6830884 | *Coccidioides posadasii* | Venezuela | Previously published |
| SRR3468051 | *Coccidioides posadasii* | Mexico | Previously published |
| SRR3468033 | *Coccidioides posadasii* | Arizona | Previously published |
| SRR3468061 | *Coccidioides posadasii* | Arizona | Previously published |
| SRR21292481 | *Coccidioides posadasii* | Washington | Previously published |
| SRR3468032 | *Coccidioides posadasii* | Arizona | Previously published |
| SRR21204708 | *Coccidioides posadasii* | Washington | Previously published |
| SRR21295064 | *Coccidioides posadasii* | Oregon | Previously published |
| SRR3468070 | *Coccidioides posadasii* | Guatemala | Previously published |
| SRR21295065 | *Coccidioides posadasii* | Oregon | Previously published |
| SRR3468062 | *Coccidioides posadasii* | Arizona | Previously published |
| SRR21204732 | *Coccidioides posadasii* | New Mexico | Previously published |
| SRR3468034 | *Coccidioides posadasii* | Arizona | Previously published |
| SRR21295068 | *Coccidioides posadasii* | Washington | Previously published |
| SRR3468057 | *Coccidioides posadasii* | Arizona | Previously published |
| SRR6830882 | *Coccidioides posadasii* | Venezuela | Previously published |
| SRR3468055 | *Coccidioides posadasii* | Arizona | Previously published |
| SRR3468053 | *Coccidioides posadasii* | Mexico | Previously published |
| SRR3468050 | *Coccidioides posadasii* | Colorado | Previously published |
| SRR3468047 | *Coccidioides posadasii* | Arizona | Previously published |
| SRR3468067 | *Coccidioides posadasii* | Guatemala | Previously published |
| SRR6830886 | *Coccidioides posadasii* | Venezuela | Previously published |
| SRR3468025 | *Coccidioides posadasii* | Arizona | Previously published |
| SRR21204721 | *Coccidioides posadasii* | Oregon | Previously published |
| SAMN57541301 | *Coccidioides posadasii* | Arizona | This Study |
| SRR3468065 | *Coccidioides posadasii* | Mexico | Previously published |
| SRR6830881 | *Coccidioides posadasii* | Venezuela | Previously published |
| SRR6830888 | *Coccidioides posadasii* | Venezuela | Previously published |
| SRR21204676 | *Coccidioides posadasii* | Oregon | Previously published |
| SAMN57541302 | *Coccidioides posadasii* | Texas | This Study |
| SAMN52167856 | *Coccidioides posadasii* | Nevada | This Study |
| SAMN57541303 | *Coccidioides posadasii* | Arizona | This Study |
| SAMN57541306 | *Coccidioides posadasii* | California | This Study |
| SAMN57541308 | *Coccidioides posadasii* | Texas | This Study |
| SAMN57541309 | *Coccidioides posadasii* | Texas | This Study |
| SAMN57541310 | *Coccidioides posadasii* | Oregon | This Study |
| SAMN52167864 | *Coccidioides posadasii* | Utah | This Study |
| SAMN57541313 | *Coccidioides posadasii* | Texas | This Study |
| SAMN57541315 | *Coccidioides posadasii* | Texas | This Study |
| SAMN57541317 | *Coccidioides posadasii* | Texas | This Study |
| SAMN57541319 | *Coccidioides posadasii* | Arizona | This Study |
| SAMN52167850 | *Coccidioides posadasii* | Colorado | This Study |
| SAMN57541321 | *Coccidioides posadasii* | Arizona | This Study |
| SAMN52167867 | *Coccidioides posadasii* | Utah | This Study |
| SAMN52167866 | *Coccidioides posadasii* | Utah | This Study |
| SAMN57541322 | *Coccidioides posadasii* | Arizona | This Study |
| SAMN57541324 | *Coccidioides posadasii* | Arizona | This Study |
| SAMN57541325 | *Coccidioides posadasii* | Texas | This Study |
| SAMN57541327 | *Coccidioides posadasii* | Arizona | This Study |
| SAMN57541328 | *Coccidioides posadasii* | Texas | This Study |
| SAMN57541329 | *Coccidioides posadasii* | Arizona | This Study |
| SAMN57541330 | *Coccidioides posadasii* | Arizona | This Study |
| SAMN52167862 | *Coccidioides posadasii* | Utah | This Study |
| SAMN57541331 | *Coccidioides posadasii* | Oklahoma | This Study |
| SAMN57541332 | *Coccidioides posadasii* | California | This Study |
| SAMN57541333 | *Coccidioides posadasii* | Texas | This Study |
| SAMN57541335 | *Coccidioides posadasii* | Arizona | This Study |
| SAMN57541336 | *Coccidioides posadasii* | Texas | This Study |
| SAMN57541338 | *Coccidioides posadasii* | Texas | This Study |
| SAMN57541344 | *Coccidioides posadasii* | Texas | This Study |
| SAMN57541345 | *Coccidioides posadasii* | Texas | This Study |
| SAMN52167848 | *Coccidioides posadasii* | Colorado | This Study |
| SAMN57541347 | *Coccidioides posadasii* | Arizona | This Study |
| SAMN52167858 | *Coccidioides posadasii* | Nevada | This Study |
| SAMN52167849 | *Coccidioides posadasii* | Colorado | This Study |
| SAMN57541348 | *Coccidioides posadasii* | California | This Study |
| SAMN57541349 | *Coccidioides posadasii* | Texas | This Study |
| SAMN57541350 | *Coccidioides posadasii* | Arizona | This Study |
| SAMN57541352 | *Coccidioides posadasii* | Arizona | This Study |
| SAMN57541354 | *Coccidioides posadasii* | Arizona | This Study |
| SAMN57541355 | *Coccidioides posadasii* | Oregon | This Study |
| SAMN57541358 | *Coccidioides posadasii* | Texas | This Study |
| SAMN57541359 | *Coccidioides posadasii* | Texas | This Study |
| SAMN57541360 | *Coccidioides posadasii* | Arizona | This Study |
| SAMN57541361 | *Coccidioides posadasii* | Arizona | This Study |
| SAMN52167851 | *Coccidioides posadasii* | Colorado | This Study |
| SAMN57541362 | *Coccidioides posadasii* | Arizona | This Study |
| SAMN57541367 | *Coccidioides posadasii* | Arizona | This Study |
| SAMN57541368 | *Coccidioides posadasii* | Texas | This Study |
| SAMN57541369 | *Coccidioides posadasii* | Arizona | This Study |
| SAMN57541371 | *Coccidioides posadasii* | Arizona | This Study |
| SAMN57541372 | *Coccidioides posadasii* | Washington | This Study |
| SAMN57541373 | *Coccidioides posadasii* | California | This Study |
| SAMN57541374 | *Coccidioides posadasii* | California | This Study |
| SAMN57541376 | *Coccidioides posadasii* | Arizona | This Study |
| SAMN52167846 | *Coccidioides posadasii* | California | This Study |
| SAMN52167873 | *Coccidioides posadasii* | Utah | This Study |
| SAMN57541379 | *Coccidioides posadasii* | Texas | This Study |
| SAMN57541380 | *Coccidioides posadasii* | Texas | This Study |
| SAMN57541382 | *Coccidioides posadasii* | Washington | This Study |
| SAMN57541383 | *Coccidioides posadasii* | Arizona | This Study |
| SAMN57541384 | *Coccidioides posadasii* | Texas | This Study |
| SAMN57541385 | *Coccidioides posadasii* | Arizona | This Study |
| SAMN57541386 | *Coccidioides posadasii* | Arizona | This Study |
| SAMN52167872 | *Coccidioides posadasii* | Utah | This Study |
| SAMN57541387 | *Coccidioides posadasii* | Arizona | This Study |
| SAMN57541388 | *Coccidioides posadasii* | Arizona | This Study |
| SAMN57541389 | *Coccidioides posadasii* | Texas | This Study |
| SAMN57541391 | *Coccidioides posadasii* | Texas | This Study |
| SAMN57541393 | *Coccidioides posadasii* | Texas | This Study |
| SAMN52167854 | *Coccidioides posadasii* | Colorado | This Study |
| SAMN52167860 | *Coccidioides posadasii* | Utah | This Study |
| SAMN57541395 | *Coccidioides posadasii* | Arizona | This Study |
| SAMN52167859 | *Coccidioides posadasii* | Utah | This Study |
| SAMN57541397 | *Coccidioides posadasii* | Arizona | This Study |
| SAMN57541399 | *Coccidioides posadasii* | Arizona | This Study |
| SAMN57541401 | *Coccidioides posadasii* | Texas | This Study |
| SAMN57541402 | *Coccidioides posadasii* | Texas | This Study |
| SAMN57541404 | *Coccidioides posadasii* | Arizona | This Study |
| SAMN57541405 | *Coccidioides posadasii* | Arizona | This Study |
| SAMN52167855 | *Coccidioides posadasii* | Nevada | This Study |
| SAMN57541406 | *Coccidioides posadasii* | Arizona | This Study |
| SAMN57541408 | *Coccidioides posadasii* | Arizona | This Study |
| SAMN57541410 | *Coccidioides posadasii* | Arizona | This Study |
| SAMN57541413 | *Coccidioides posadasii* | Arizona | This Study |
| SAMN57541417 | *Coccidioides posadasii* | Arizona | This Study |
| SAMN52167861 | *Coccidioides posadasii* | Utah | This Study |
| SAMN57541418 | *Coccidioides posadasii* | Texas | This Study |
| SAMN57541419 | *Coccidioides posadasii* | Texas | This Study |
| SAMN57541420 | *Coccidioides posadasii* | Arizona | This Study |
| SAMN57541421 | *Coccidioides posadasii* | Arizona | This Study |
| SRR25495556 | *Coccidioides posadasii* | Brazil | Previously published |
| SRR25495557 | *Coccidioides posadasii* | Brazil | Previously published |
| SRR25495558 | *Coccidioides posadasii* | Brazil | Previously published |
| SRR25495559 | *Coccidioides posadasii* | Brazil | Previously published |
| SRR25495560 | *Coccidioides posadasii* | Brazil | Previously published |
| SRR25495561 | *Coccidioides posadasii* | Brazil | Previously published |
| SRR25495562 | *Coccidioides posadasii* | Brazil | Previously published |
| SRR25495563 | *Coccidioides posadasii* | Brazil | Previously published |
| SRR25495564 | *Coccidioides posadasii* | Brazil | Previously published |
| SRR25495565 | *Coccidioides posadasii* | Brazil | Previously published |
| SRR25495566 | *Coccidioides posadasii* | Brazil | Previously published |
| SRR25495567 | *Coccidioides posadasii* | Brazil | Previously published |
| SRR25495568 | *Coccidioides posadasii* | Brazil | Previously published |

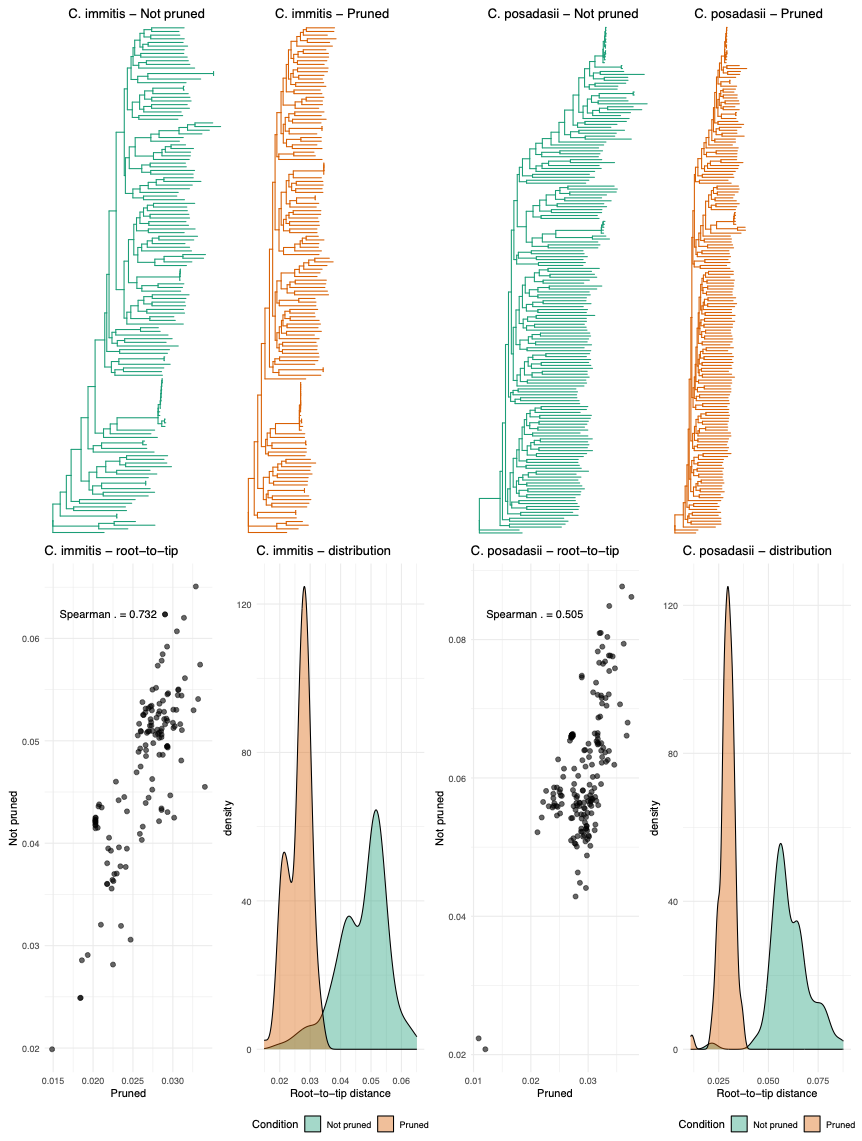
**Figure S1.** Comparison of phylogenetic branch lengths inferred from linkage disequilibrium (LD)-pruned and unpruned datasets for *Coccidioides immitis* and *C. posadasii*. Maximum-likelihood trees reconstructed with IQ-TREE are shown for the unpruned and LD-pruned datasets (top panels). Corresponding root-to-tip distances were positively correlated between datasets, with a stronger association for *C. immitis* (Spearman’s ρ = 0.732) than for *C. posadasii* (Spearman’s ρ = 0.505; bottom scatterplots). Density distributions show that root-to-tip distances were systematically greater in the unpruned datasets, indicating that retaining linked SNPs increased the inferred branch-length scale.

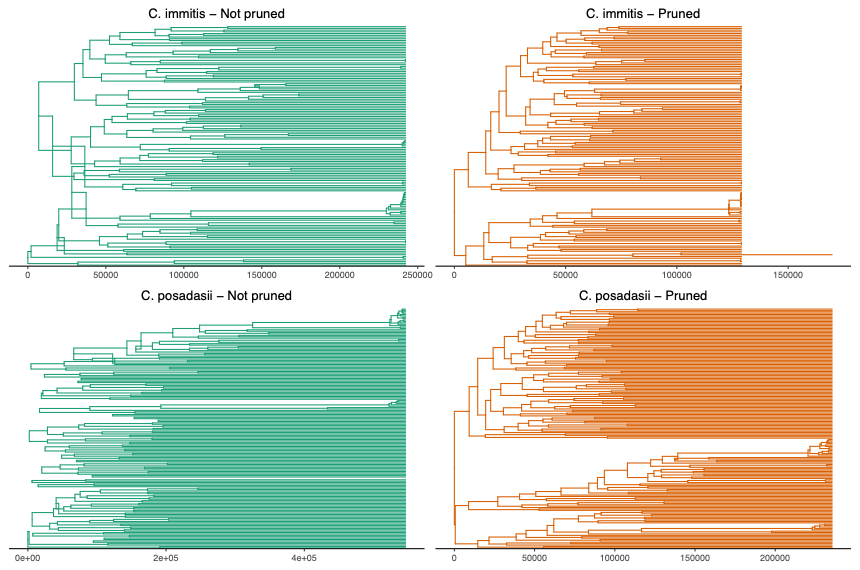
**Figure S2.** Time-calibrated phylogenetic trees inferred from linkage disequilibrium (LD)-pruned and unpruned datasets for *Coccidioides immitis* and *C. posadasii*. Trees were reconstructed using TreeTime, with branch lengths expressed in years before present. For both species, analyses of the unpruned datasets produced longer branches and substantially older divergence-time estimates than analyses of the LD-pruned datasets. This systematic shift indicates that retaining correlated SNPs in linkage disequilibrium inflates the inferred amount of molecular change and, consequently, the estimated evolutionary timescale.

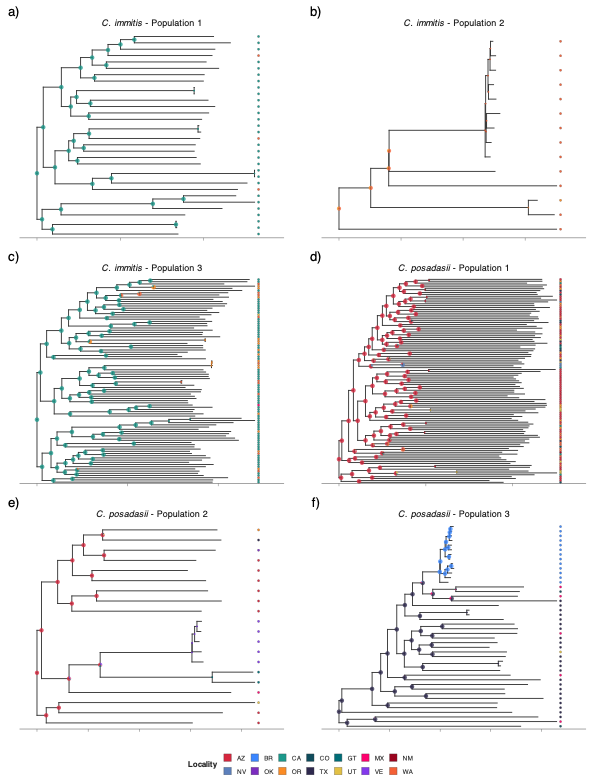
**Figure S3.** Historic patterns of gene flow from ancestral state reconstruction. For each genetic population, a maximum likelihood phylogeny with a heat map to the right of the tree indicating the state or region where each isolate was collected. Pie charts at internal nodes represent the probability that the ancestor occurred in each inferred geographic location.
